# Acceptability and perspectives on clinic-based urine tenofovir testing for antiretroviral therapy adherence monitoring: qualitative findings from a randomized controlled trial in South Africa

**DOI:** 10.1101/2025.03.07.25323040

**Authors:** Ashley R. Bardon, Makhosazane Zondi, Jane M. Simoni, Kwena Tlhaku, Pedzisai Munatsi, Nomfundo Bhengu, Elex Hill, Mlungisi Khanyile, Monica Gandhi, Jienchi Dorward, Nigel Garrett, Paul K. Drain

## Abstract

Real-time, urine tenofovir testing may allow for clinic-based monitoring of adherence to antiretroviral therapy (ART). We aimed to assess (1) the acceptability of monthly point-of-care urine tenofovir testing over the first five months following ART initiation and (2) perspectives on the implementation of point-of-care urine tenofovir testing among people living with HIV (PLWH) and healthcare providers participating in a randomized controlled trial which used the urine test in South Africa. We conducted in-depth interviews with 20 PLWH six-months post-ART initiation and with eight healthcare providers. We assessed the acceptability (using constructs from the Theoretical Framework of Acceptability), appropriateness, feasibility, and willingness to use the point-of-care urine tenofovir test, as well as participants’ preferred form of adherence monitoring and perspectives on differentiated implementation strategies. Participants found monthly point-of-care tenofovir testing highly acceptable, preferrable to self-reported adherence measures, appropriate for this population, and potentially feasible to integrate with standard-of-care ART monitoring. Participants’ overall acceptability of routine urine tenofovir testing was shaped by experiences and perceptions that shaped their overall acceptability. Routine urine tenofovir testing was well-liked, perceived to be low-burden with few opportunity costs, and perceived to have several positive effects. These included encouraging consistent ART adherence, strong client-provider relationship and communications, and accurate self-reporting of adherence. Participants’ desire to impress and build trust with their provider motivated them to take their ART daily to achieve a positive adherence test result at each clinic visit. Overall, point-of-care urine tenofovir testing may be an acceptable and beneficial tool for motivating optimal adherence, improving ART adherence monitoring, and strengthening client-provider relations.

## INTRODUCTION

South Africa has the largest HIV epidemic in the world but has significant gaps in HIV treatment coverage and viral suppression [1]. Progress in achieving the 95-95-95 goals may be limited by achieving and sustaining optimal adherence during the first six months of treatment, a crucial period when people living with HIV (PLWH) begin to establish behaviors [2]. About half of PLWH initiating antiretroviral therapy (ART) in South Africa experience adherence challenges at least once in the first six months of treatment, and an estimated 30% of PLWH disengage from care within the first six months of treatment [3–7]. Developing and sustaining early, optimal adherence behaviors and preventing disengagement from care through targeted, person-centered interventions will be critical to ending the HIV/AIDS epidemic in South Africa [8].

Early interventions targeting PLWH who are experiencing adherence challenges or other barriers to care are particularly needed to prevent adverse clinical outcomes [8]. Clinicians rely on subjective measures to identify adherence challenges, such as self-reported behaviors, which may be inaccurate and unreliable for informing clinical decisions and counseling support [9–15]. The need for adherence monitoring is particularly relevant in the first months after ART initiation, when the viral load has yet to suppress and can’t be used as a measure of adherence. Point-of-care (POC) urine tenofovir testing is a new development in therapeutic drug monitoring that can measure recent (prior 2-4 days) ART adherence in real time, allowing for prompt identification of adherence challenges and immediate interventions for adherence support [16–20]. POC urine tenofovir testing also has the potential to predict future loss to follow-up, viremia, and drug resistance [21–24] and may motivate PLWH to improve their ART adherence [13,19,25–27]. We conducted the Simplifying TREAtment and Monitoring for HIV (STREAM HIV) trial to test the effect of a combined intervention of monthly POC urine tenofovir testing for the first five months of HIV treatment and POC HIV viral load testing at six and 12 months in comparison to standard-of-care HIV treatment services on the outcomes of ART adherence, retention in care, and virologic suppression [28].

Some studies have found the POC urine tenofovir test to be generally acceptable to PLWH and healthcare providers and appropriate for monitoring ART adherence [20,25,26,29]. However, some PLWH and healthcare providers have had neutral or negative perceptions of the test, expressing uncertainty about the test’s overall utility since it can only detect recent adherence and concerns about the impact of drug-level testing on client-provider relationships, particularly clients’ perceptions of mistrust from their provider [19,26,29]. These findings may indicate that POC urine tenofovir testing is suitable and acceptable for certain populations and settings, although it may not be universally applicable. Still, little is known about how a longitudinal intervention of monthly POC urine tenofovir testing will be perceived and accepted by PLWH and healthcare providers and how the test will affect the client-provider relationship and client’s behaviors, such as ART adherence and retention in care. In this qualitative sub-study of the STREAM HIV trial, we sought to understand the acceptability of a monthly POC urine tenofovir testing intervention over the first five months of treatment among PLWH and healthcare providers in South Africa. To inform future interventions of POC urine tenofovir testing, we also collected perspectives from PLWH and healthcare providers on differentiated approaches for implementing POC urine tenofovir testing.

## METHODS

### Study Design and Setting

We conducted a qualitative sub-study nested within the STREAM HIV trial (ClinicalTrials.gov: NCT04341779) among PLWH and healthcare providers participating in the trial intervention procedures [28]. STREAM HIV is a two-arm, open-label randomized controlled trial testing the effect of a combined intervention of monthly POC urine tenofovir testing for the first five months of HIV treatment, along with routine POC HIV viral load testing at six and 12 months, in comparison to standard-of-care HIV treatment services. Trial outcomes include ART adherence, retention in care, and viral suppression at 12 and 18 months. More details about the STREAM HIV trial can be found in the published protocol [28].

The STREAM HIV trial was conducted at two sites in KwaZulu-Natal, South Africa: (1) the Centre for the AIDS Programme of Research in South Africa (CAPRISA) eThekwini Clinical Research Site (CRS) in urban eThekwini (Durban), which is located adjacent to a large public HIV clinic (Prince Cyril Zulu Communicable Disease Centre) and the Lancers Road Clinic, a primary care clinic; and (2) the CAPRISA Vulindlela CRS in rural uMgungundlovu District, which is adjacent to a public primary healthcare clinic (Mafakatini Clinic). Both research sites are adjacent to large, public clinics serving a high volume of PLWH receiving ART.

#### Study Population

Individuals were eligible to participate in the STREAM HIV trial if they were ≥16 years old, living with HIV, initiating tenofovir disoproxil fumarate (TDF)-based ART, not taking any ART regimen in the prior month, and were willing and able to provide written informed consent. We enrolled two populations in this qualitative sub-study: intervention arm participants who received POC TFV testing (“PLWH”) and healthcare providers who provided care to trial participants (“providers”). Trial participants were eligible to participate in this qualitative sub-study if they were enrolled in STREAM HIV, were returning for a six-month clinic visit, had been randomly assigned to the intervention arm, and were willing and able to provide written informed consent. We purposively sampled a subset of intervention arm participants to achieve a diverse sample in terms of age, sex, and adherence test results (detectable tenofovir test results at all visits or one or more undetectable tenofovir test results). PLWH were only included from the eThekwini CRS.

Healthcare providers were eligible to participate in the qualitative sub-study if they had provided care for participants in the intervention arm of the STREAM HIV trial and were willing and able to provide written informed consent. We sampled all providers who were eligible and willing to participate after about half of STREAM HIV participants had reached the six-month endpoint. We included healthcare providers from both the eThekwini CRS and the Vulindlela CRS.

#### Study Procedures and Data Collection

We conducted semi-structured in-depth interviews (IDIs) with participants randomized to the intervention arm of the STREAM HIV trial at their six-month follow-up visit. Participants in the intervention arm received POC urine tenofovir testing at each monthly follow-up visit for the first five months. The POC urine tenofovir test was performed by a research nurse in a private exam room in the presence of the participant (processing time of ∼5 minutes), and the nurse and participant reviewed the test results together. We recruited intervention arm participants for IDIs in person at their six-month follow-up visit, and we recruited healthcare providers for an IDI via email. No intervention arm participants or healthcare providers refused to participate in an IDI or withdrew their participation.

All IDIs were conducted in a private room in the participants’ preferred language (English or isiZulu). Each IDI participant was interviewed by a female research assistant (ARB, NB, MZ, KT) who was not involved in patient care, and another member of the research team was present during each IDI to support data collection and audio recording. All interviewers had Master’s degrees with formal training and experience in qualitative research methods. PLWH were compensated 100 South African Rand (∼6 U.S. dollars) for their participation in the IDI; healthcare providers were interviewed during their working hours and did not receive additional compensation. At the beginning of each IDI, the interviewer introduced herself and explained the purpose of the IDI. The interviewer or another member of the research team took notes during each IDI for future reference. All IDIs were audio recorded, transcribed, translated to English if necessary, and checked for accuracy against the audio recordings by the research team. Demographic and clinical information was collected from PLWH at enrollment, and demographic information was collected from healthcare providers at the time of their IDI.

#### Instrument Design

We developed one interview guide for PLWH (Supplement 1) and one for providers (Supplement 2). The participant interview guides were developed to capture the following domains: comprehension of the intervention; understanding of the rationale for POC urine tenofovir testing; attitudes about the intervention; experiences with the intervention; perceived effect of the intervention on self-reported adherence, client-provider relationship, ART adherence, and other behaviors; preferred ART adherence monitoring approach (POC urine tenofovir testing versus self-reporting adherence) and perspectives on other ways POC urine tenofovir testing can be implemented (i.e., who would benefit from testing, where it could be implemented, when it could be implemented, and who could administer/perform the test).

Provider interview guides included the following domains: comprehension of the intervention, understanding of the rationale for POC urine tenofovir testing, attitudes about the intervention, experiences with the intervention, approach to communicating POC urine tenofovir test results to participants, barriers and facilitators to communicating test results, perception of participants’ understanding of and reaction to test results, perceived feasibility of the intervention, and perspectives on other ways POC urine tenofovir testing can be implemented (i.e., who would benefit from testing, where it could be implemented, when it could be implemented, and who could administer/perform the test).

#### Data Analyses

We (ARB, NB) created one codebook for PLWH and one for healthcare providers, which were deductively developed based on domains of interest from the interview guides. We also applied an inductive approach during the coding process to add new codes for topics that were discussed outside of the domains of interest. Each transcript was independently coded by two coders (ARB, NB), compared to identify discrepancies, and recoded when necessary. After consensus that data saturation was achieved and all coding discrepancies were resolved between the two coders, we performed thematic analyses to identify and describe emergent themes. Most themes overlapped with constructs from the Theoretical Framework of Acceptability (TFA) (affective attitude, burden, intervention coherence, effectiveness, opportunity costs); therefore, we created a thematic map of all the themes related to acceptability using this framework [30]. We also mapped themes related to implementation of POC urine tenofovir testing using the Differentiated Service Delivery elements (context, clinical characteristics, specific populations) and building blocks (who, what, when, where) [31]. Additional themes were described for preferences for adherence monitoring, willingness to implement the intervention, and perceptions about the appropriateness and feasibility of the intervention. All transcripts were stored, coded, and analyzed using Dedoose software (SocioCultural Research Consultants, LLC, Los Angeles, CA, USA). We also used descriptive statistics to present demographic and baseline clinical characteristics (only PLWH) of participants included in this qualitative study.

#### Ethical Considerations

The STREAM HIV trial received ethical approvals from the University of KwaZulu-Natal Biomedical Research Ethics Committee (BREC/00000833/2019), the University of Washington Human Subjects Division (STUDY00007544), and the National Institutes of Health Division of AIDS Regulatory Support Center (DAIDS-ES ID #38509). All participants provided written informed consent to participate in qualitative interviews, as well as the trial.

## RESULTS

We enrolled 28 participants in this qualitative sub-study of the STREAM HIV trial; 20 PLWH from the intervention arm, and eight healthcare providers (**Table 1**). The median age was 38.5 years for PLWH and 36.5 years for providers. Half (50%, 10/20) of PLWH and 88% (7/8) of providers were female. Among PLWH, most did not know their primary partner’s HIV status (66.7%, 12/18), and half had a CD4 count <200 cells/mm^3^ (50%, 10/20). Three PLWH (15%) had received one undetectable tenofovir test result, and 85% (17/20) had received only detectable tenofovir results. The providers included an enrolled nurse assistant, research nurses, and clinicians. The median duration of IDIs was 50 minutes (IQR 44-55 minutes) for PLWH and 45 minutes (IQR 40-60 minutes) for providers.

**Table 1.**
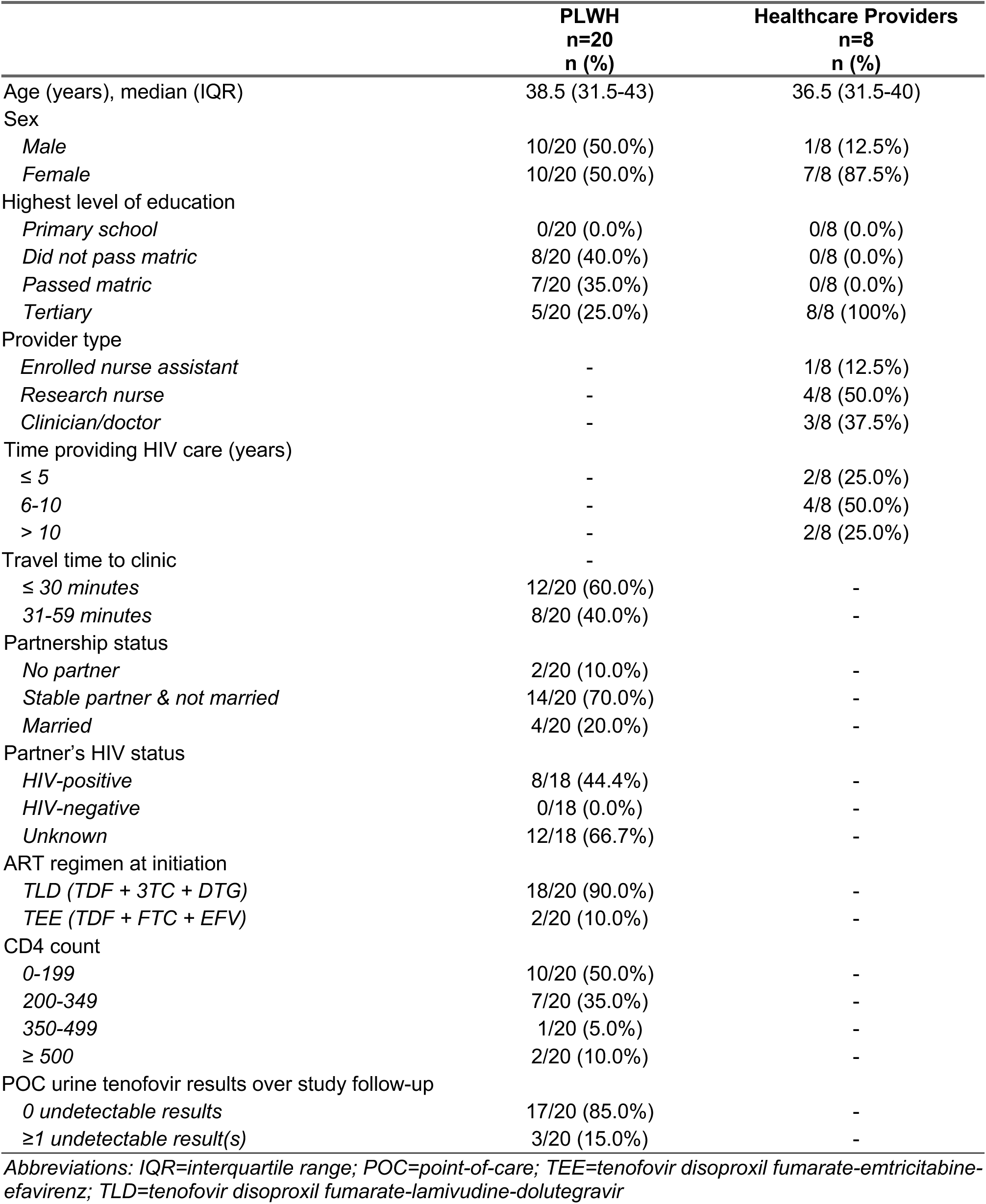
Baseline characteristics of qualitative study participants, N=28.

### Acceptability of monthly POC urine tenofovir testing

Overall, PLWH and providers perceived the monthly POC urine tenofovir testing intervention as highly acceptable. We mapped each theme to relevant constructs from the TFA. We summarize emergent themes by TFA constructs in **Figure 1** and illustrate these themes with supporting quotes within each section and in **Table 2**.

**Figure 1.**
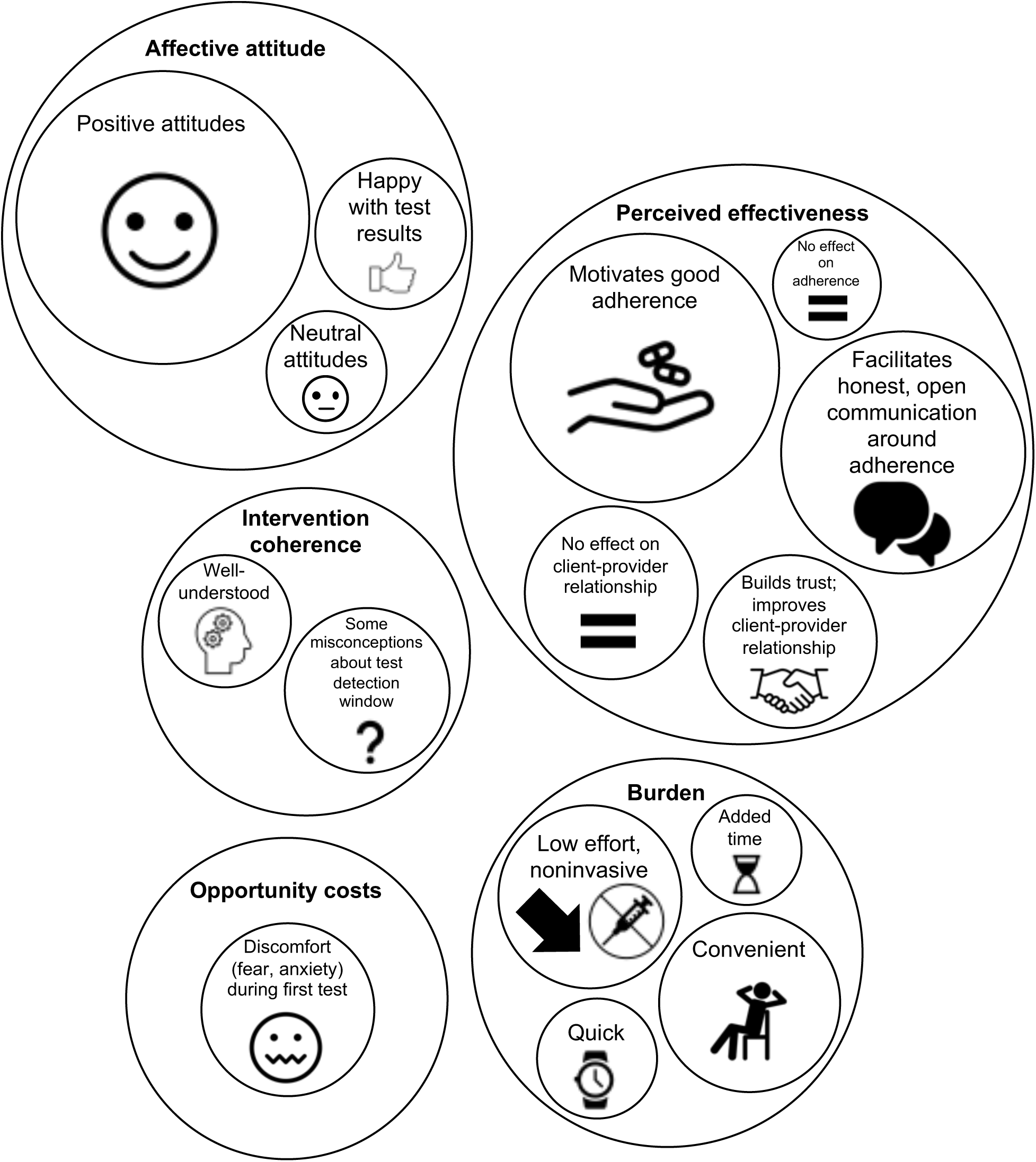
Thematic map of acceptability of monthly point-of-care urine tenofovir testing, organized by acceptability constructs.

### Affective attitude

The majority of PLWH liked the intervention, and several stated that seeing their test results made them feel happy. PLWH expressed that adherence testing made them feel supported by their providers and demonstrated that the providers cared about the wellbeing of their clients. PLWH liked the intervention because it held them accountable for their adherence, it facilitated open and honest adherence discussions with their provider(s), they were recognized for their good adherence, and they were included in the testing process. Providers also described observing some PLWH as eager to use the adherence test.

*“I liked it because I saw that it meant I’ll be getting help.” (Intervention participant, female, 40-49 years old)*

*“It showed positive, one line. And it made me happy, and I had that closure that ‘oh! I’m doing it the proper way.’” (Intervention participant, female, 20-29 years old)*

The remainder of PLWH expressed neutral attitudes about the intervention.

*“It’s okay if it’s part of the study to help other people, but for me personally, it does nothing. I don’t gain nor lose anything due to fact that I know how my pills work.” (Intervention participant, male, 40-49 years old)*

All of the providers had positive attitudes about the intervention.

*“I think it’s a good thing. I think it’s beneficial.” (Healthcare provider, female, urban site)*

### Perceived effectiveness (and secondary benefits)

Overall, most participants perceived the intervention to have many positive effects and benefits for PLWH and providers. All providers and nearly all PLWH perceived the intervention as having a positive impact on participants’ overall adherence. The test was motivational to PLWH to take their ART daily and improve their adherence.

*“It had positive impact because it encourages me to always remember my pills not just when the next date to [the clinic] is near, I’m not doing it for [the clinic], I’m doing it for myself.” (Intervention participant, female, 30-39 years old)*

PLWH often thought about their upcoming adherence test when they were at home, which served as a daily reminder to take their ART. Several PLWH believed the test was able to detect adherence over an entire month, which prompted them to take their ART as prescribed daily. Two participants understood that the test’s ability to only detect recent dosing could influence some PLWH to take a pill before an upcoming adherence test to manipulate the test results, though no participants reported attempting to manipulate the test in that way.

*“I would say that the test played some sort of an alarm to me. By saying an alarm, I mean that, I knew if I miss a day or two of pills there will be something that will tell that I wasn’t honest in taking the treatment. So that’s the role that the tests played and was sort of like a motivator because I knew that it will be evident if I took them and if I didn’t take them in three or four days. So I was happy that these tests made me to comply to taking my pills constantly.” (Intervention participant, male, 30-39 years old)*

Some PLWH felt internally incentivized to improve their adherence and take control of their health, but many described their motivation to improve ART adherence was driven by the desire to have a ‘good’ adherence test result to provide evidence of their adherence, impress their provider, build trust with their provider(s), and receive recognition and praise for their effort. PLWH were also motivated by the fear of being exposed if their adherence test were to reveal undetectable tenofovir, which they perceived would be a disappointment to their provider. One PLWH believed that their provider may react poorly and ‘fight’ with them if they were to receive an undetectable tenofovir test result. A couple of PLWH perceived the intervention as having no effect on their adherence, but no participants perceived the intervention as having a negative effect on adherence.

*“Yes, it had an impact because it encouraged me to take the pills the way I am supposed to. The way the nurses were so welcoming, not just one of them but all of them it made it seem like we were a team so that also encouraged me to take the pills right. Not just one person, sometimes I would find a different nurse from the one that I saw previously, but they are just the same it really gave peace and spend most of the time with them.” (Intervention participant, male, 30-39 years old)*

*“…it will strengthen adherence, because with them, knowing that you’re going to be testing, they obviously don’t want to come to you, and then have a bad result. So, I think it will also motivate them a little bit to say, because we’re going to be testing for this, they would be a little bit more motivated to, or determined I’d say, to take their treatment so their results are good when they do come to us. So, I mean, psychologically, it may also have an effect and help with adherence.” (Healthcare provider, female, urban site)*

The intervention was also perceived to be more effective than standard-of-care for providing early evidence of adherence (before participants’ first viral load test) and prompting appropriate adherence interventions.

*“I think that [tenofovir testing is] a good way to detect whether the patient, the participant, really has, you know, some issues early. Like it’s an earlier stage than having to wait for the viral load at six months, so that’s like…cause from the first 5 months, you have to just hear from what the participant tells you. If they say they take their medication, then you can’t dispute that. So for the POC arm, it’s easier to see whether they are really missing the doses, so I think that’s like early detection of defaulting and all that.” (Healthcare provider, female, urban site)*

PLWH admitted that they were not always honest with their provider and that they would probably be dishonest or exaggerate their adherence if they had not received objective adherence testing. Most PLWH perceived adherence testing as having a positive impact on their self-reported adherence and other behaviors because they believed their adherence would be exposed through the test. As a result, their openness about adherence challenges helped facilitate additional conversations and a more targeted discussion around adherence.

*“I was telling the truth because urine was even confirming for me.” (Intervention participant, male, 30-39 years old)*

Providers echoed many of PLWH’ perspectives, as they also perceived the intervention as beneficial for improving discussions around adherence.

*“I think between the two arms, in the intervention one, they get to have something that makes them to take their medication because they’re like it’s going to be seen. The others, they don’t know about the test, so whether they tell you that I’m taking it right, it’s not like the intervention arm where they know maybe I’ll be caught. So I’ll just have to say I didn’t take them because there’s a plate the way you ask them before, like I normally ask them before I do the test, and then they tell you all. ‘Yes, I took my medication.’ But with this intervention ones, they tell you the honest truth, you know. ‘I forgot them maybe for five days.’ ‘No, someone took them from me.’ Or ‘I forgot. I left them at home and I was somewhere.’ So you get the real story, comparing with the ones that they’re always saying, ‘No, I’m taking them correctly.’” (Healthcare provider, female, rural site)*

Half of PLWH also emphasized that the intervention had a positive impact on the client-provider relationship. They perceived that adherence testing helped build trust between clients and providers as the test results provided evidence of their honesty. One PLWH felt as though they were on a team with their providers working towards a common goal, while another PLWH explained that they had more time to get to know their provider while the adherence test was processing. Several PLWH were also appreciative of being included in the testing process, as providers performed the test in real-time in the presence of the participant and also encouraged participants to interpret their own test results. The other half of participants perceived the intervention as having no effect on their relationship with their provider.

*“[Our relationship] was going to differ [if there was no adherence testing] because there was going to be nothing confirming that the pill is indeed in my body. No further questions were going to be asked other than asking if you’re taking pills and get another pill then leave like it happens in other clinics, people complain about that.” (Intervention participant, female, 30-39 years old)*

### Intervention coherence

The intervention procedures and rationale were well-understood by PLWH and providers with the exception of two PLWH who had forgotten how to interpret the test results by the time of their IDI at the six-month visit. However, the test’s limited ability to detect only recent adherence was not fully understood by all PLWH. PLWH had several misconceptions about the test, including i) the test could detect adherence over an entire month, ii) it could detect if a pill was taken late one or more days over the past month, iii) the test could help uncover other issues interfering with drug absorption, and drugs or iv) alcohol could interfere with the test results. Providers may have partially contributed to the misconceptions by not correcting intervention participants’ misunderstanding and withholding information about the test’s limitations, particularly with regards to the test’s detection window for tenofovir.

*“So because we don’t explain to them that if they don’t take the treatment four days, if you didn’t take the treatment four days ago. Because if you say that, then maybe they’ll make sure that these four days, you’re taking your treatment before you go to the clinic. So we don’t give that information unnecessarily. So maybe this person has not been taking treatment like they’re supposed to. Yeah. They have been taking treatment, but maybe at 9:00, maybe at 8:00 sometimes. So now you’re not sure what’s going to come then. I think that’s the problem. And then when it comes back positive, they’re relieved and like, ‘yeah.’” (Healthcare provider, female, urban site)*

### Burden

Overall, participants found the intervention to have little to no burden to PLWH and providers. POC urine tenofovir testing was summarized as quick, easy, convenient, and required little to no effort.

*“It was comfortable because they put the test in front of me to see while we do the test and wait for the results, so everything was done out in the open. I think it’s more convenient if they get them at the same time cause it takes only a few minutes. So, it’s more convenient if they hear them [at] the [same] time they’re there than hearing them any other way, and you get to see them for yourself. There’s no waiting for the results to come back from the lab.” (Intervention participant, female, 20-29 years old)*

*“I think it’s just easy. It’s convenient because the test is right here. You do it while the participant is here. It’s just, you know, I haven’t seen anything that would cause me to be like inconvenienced. It doesn’t take long.” (Healthcare provider, female, rural site)*

PLWH were unable to draw comparisons to standard HIV care in terms of the intervention’s burden, because they had not experienced care without the intervention. However, a few PLWH believed that the intervention may add some extra time to each follow-up visit. Providers, on the other hand, believed that adherence testing did not add any additional time to participants’ follow-up visits since they were able to perform other tasks while participants were collecting urine or while the test was processing for five minutes. Some PLWH and providers also felt that urine testing is acceptable since it’s noninvasive compared to other tests that require needles and not burdensome to clients since it requires only a small amount of urine.

*“I think urine is quite an acceptable means of testing. It’s non-invasive. There’s no needle stick. If I was a patient, I would find that method acceptable. It’s comfortable. It’s not time-consuming. So I think all those aspects of it, I’m very in favor of it therefore being a part of standard of care.” (Healthcare provider, female, urban site)*

### Opportunity costs

Many PLWH described feeling uncomfortable, worried, anxious, or fearful during their first adherence test, even if they were confident that their adherence was good over the prior month. Generally, these feelings dissipated by subsequent tests except when their adherence was poor in the prior month.

*“In the first visit I was afraid. But as time went on, I got used to it.” (Intervention participant, female, 20-29 years old)*

Providers also perceived PLWH to be slightly uncomfortable during their first adherence test but comfortable during subsequent tests.

*“I think for the first time, they’ll be anxious. They’ll like stand beside you and wait for the results. But once they’ve gotten used to it, they don’t…they’re just neutral.” (Healthcare provider, female, urban site)*

#### Preferences and willingness to use

When PLWH were asked about their preferred form of adherence monitoring (POC urine tenofovir testing or self-reported adherence), nearly all stated that they would prefer POC urine tenofovir testing.

*“I think I would prefer the test to be used because it will encourage me to take it, because it’s not easy to take them to be honest. Taking pills every day is a story. So, for me it helped me because I thought it will be hard on me, but these tests motivated me because I know it will show.” (Intervention participant, female, 30-39 years old)*

However, one PLWH was neutral about their preference and only stipulated that they would choose POC urine tenofovir testing if it were quick, low cost, and cost-effective.

Similarly, all providers were very willing to use POC urine tenofovir testing for all PLWH, as they perceived the intervention as very acceptable and effective after their experience with it, though they had some skepticism about how willing providers would be to implement the test in other settings.

*“Yeah, I would definitely be willing. As I’ve said, I think it just gives you a better picture of adherence so that you know how to better support and counsel the patient or participant. And I think if you do that early on, so it would just help establish them for a good, lifelong journey with their ART.” (Healthcare provider, female, urban site)*

#### Appropriateness

The majority of participants perceived that the intervention was appropriate for their own care and treatment as well as for all PLWH in South Africa, particularly for the first five months of treatment when PLWH are beginning to establish adherence behaviors. One PLWH perceived the intervention as having little utility or value for their own care and for other PLWH who don’t require additional adherence support. All providers perceived the intervention to be appropriate for the population included in this study.

*“I think it can help many people nationally here in South Africa because in other clinics if it’s your date you just go there ask your card, show your appointment card and they give you your treatment and go, so whether you use the one they gave you before they don’t see that because there’s no tests done on their visit just a follow-up of what has been done and to see if it’s happening or not. So, if this kind of test can be implemented to all clinics for people using treatment, I think it can have much influence and things can change a lot on what the situation can be, because people can keep their viral load down and not going up because they’ll use their treatment correctly.” (Intervention participant, male, 40-49 years old)*

#### Feasibility

Overall, the intervention was perceived by providers to be very feasible to implement in standard-of-care, though they predicted several implementation barriers that would need to be addressed or overcome to make the intervention successful. The implementation barriers included physical resource availability (functional bathrooms, urine collection containers), human resource requirements (staffing, training), acceptability from key stakeholders, costs, and time.

*“I think this is revolutionary. It would really change a lot in terms of patient flow, somewhat, because obviously we’re not doing urine testing for everybody. But yeah, we can look at an adjustment to the flow because it’s such a test that can be done by a relatively lower-level category of staff, it can be taken to scale very quickly as long as we have the kits available. And if there’s strong, concrete messaging on the interpretation of the results, it’s not very complicated to roll out.” (Healthcare provider, male, urban site)*

*“I think in a clinical trial setting, so in our [clinic], we had all the infrastructure. But from my experience previously, in like the clinic-based settings or government settings,…I think it would be a challenge to implement it just because they’re so short in time, I mean short-staffed, so they try to turn over patients quickly, and I think the staff in the clinics when I perceive it as just an additional thing to do…But I think if they are properly supported, and they can see that perhaps that would lead to better outcomes and less visits in the future because people are more compliant and doing well on their treatment, that the long-term benefit would be there, and perhaps once they’re in the routine of doing it, that it wouldn’t actually be such a big ask of them.” (Healthcare provider, female, urban site)*

#### Differentiated implementation strategies for POC urine tenofovir testing

We also explored potential target populations for POC urine tenofovir testing and strategies for its implementation. Participants suggested several innovative approaches for how adherence testing could be implemented for the greatest benefit. **Figure 2** summarizes themes from participants’ exploration into differentiated implementation POC urine tenofovir testing as they fit within the elements and building blocks of differentiated service delivery.

**Figure 2.**
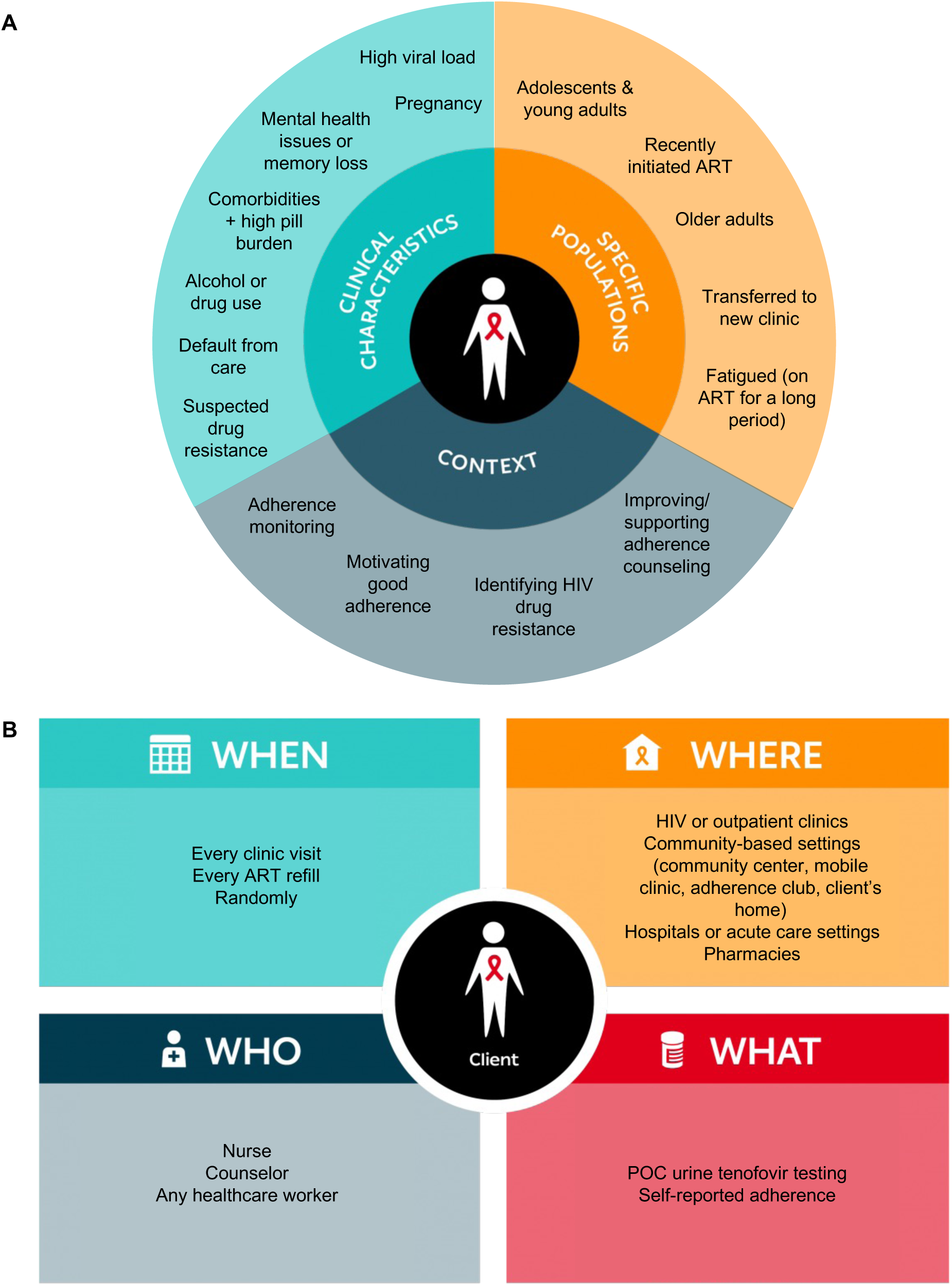
Thematic map of implementation perspectives for monthly point-of-care urine tenofovir testing, organized by elements (A) and building blocks of differentiated service delivery.

### Populations, clinical characteristics, and context

When asked about a target population for adherence testing, participants most frequently indicated that adolescents and young adults (16-30 years old) living with HIV would benefit the most from POC urine tenofovir testing, as this group experiences more adherence challenges than older populations and may be motivated by the test to improve their adherence, particularly if offered outside of the clinic setting. Participants also suggested that adherence testing could be beneficial for PLWH who had recently initiated ART, had a high viral load at a recent visit, are older, are pregnant, are experiencing mental health issues or memory loss, transferred to a new clinic, have comorbidities and a high pill burden, are fatigued by treatment, use alcohol or drugs, default from care, and have suspected HIV drug resistance. A couple of participants recommended POC urine tenofovir testing to support the identification of HIV drug resistance if results don’t match viral load test results or by establishing or confirming selective pressure prior to genotypic resistance testing. Some participants also believed that all PLWH receiving ART would benefit from regular POC urine tenofovir testing.

### When

The majority of participants felt that POC urine tenofovir testing would be most effective if it is implemented at every clinic visit or every ART refill visit, particularly for PLWH who recently initiated ART. A couple of providers suggested adherence testing could be conducted at random intervals for all PLWH to continue motivating optimal ART adherence while reducing the resources required for routine testing.

### Where

Generally, participants believed that outpatient clinics providing care for PLWH would be the most appropriate location for conducting POC urine tenofovir testing since it allows clients to be assessed for treatment side effects or other health issues at the same time. Community-based settings were also suggested as an appropriate adherence testing venue by many participants, including community centers, mobile clinics, adherence clubs, and clients’ homes. Participants believed community-based adherence testing would be most suitable for PLWH who are unable to access a clinic, PLWH who are lost to follow-up, and young PLWH who may experience other barriers to accessing a clinic. However, two PLWH had concerns about stigma and confidentiality if adherence testing is conducted at community-based settings. Hospitals, acute care settings, and pharmacies were also cited as potentially beneficial for conducting adherence testing.

### Who

When asked who should be performing POC urine tenofovir testing and delivering test results to clients, most PLWH and providers felt strongly that nurses should be performing the tests during clinic visits since they would be able to address any health or adherence concerns concurrently. Several providers also believed a counselor may be an appropriate provider for adherence testing, as they could provide adherence counseling and support during the testing process. Overall, participants felt that any healthcare workers would be able to perform adherence testing and deliver test result to clients since the test requires minimal training and resources.

## DISCUSSION

In this qualitative study among PLWH initiating ART in South Africa, we found an intervention of monthly POC urine tenofovir testing over the first five months of treatment to be highly acceptable to PLWH receiving the intervention and healthcare providers delivering the intervention. No participants expressed negative attitudes about a urine tenofovir test being incorporated into HIV treatment services, while nearly all had positive attitudes. Generally, the urine assay was perceived as having a low burden, few opportunity costs, and several positive effects and benefits to PLWH and providers. Some PLWH had misconceptions about the test’s capabilities, which may have resulted in unforeseen benefits to participants, such as motivating participants to take their ART daily between their once-monthly tests. PLWH preferred the objective adherence test over self-reporting their adherence at each visit, and providers were completely willing to use POC urine tenofovir testing for all PLWH. The urine tenofovir assay was also deemed to be appropriate for PLWH and potentially feasible for implementation as standard-of-care. Overall, the positive themes that emerged from this acceptability analysis of the urine test far outweighed the negative themes.

When we explored potential target populations and differentiated implementation strategies for POC urine tenofovir testing, participants generally perceived that the target population (PLWH initiating ART) and implementation strategy (monthly, clinic-based testing conducted by nurses in the first five months of care) in this study was appropriate and beneficial. Participants believed this intervention would be particularly beneficial for adolescents and young adults and other populations at risk of adherence challenges or defaulting on ART. They also offered several implementation strategies that should be explored for POC urine tenofovir testing, such as community-based testing, random clinic-based testing, and clinic-based testing delivered by other healthcare workers such as counselors.

To our knowledge, STREAM HIV is the first trial to evaluate a monthly POC urine tenofovir testing intervention for PLWH initiating ART, and this qualitative sub-study provides the first evidence of participants’ perceptions of and experiences with the intervention. In prior studies, POC urine tenofovir testing and other forms of drug-level testing have been found to be acceptable and perceived as valuable for improving outcomes [20,25,26,29]. Concerns about the test’s utility and impact on client-provider trust and relations have been discussed in prior studies [19,26,29] but were generally not concerns for participants in our study. In fact, nearly all participants found the monthly POC urine tenofovir testing intervention to be very useful and beneficial and to have a positive impact on trust between clients and provider and the client-provider relationship. However, the intervention’s perceived effect on motivating improved adherence may have been bolstered by some participants’ misconceptions about the test’s adherence detection window, and we may not find the same impact if participants’ misunderstandings are clarified. Additionally, while participants perceived the intervention to have a positive or neutral impact on the client-provider relationship, it likely resulted from the supportive feedback they received from the providers. Importantly, participants in this qualitative study perceived the intervention to have an overall positive impact on adherence. As access to new non-TDF-based regimens for HIV treatment expand, POC urine tenofovir testing may still be relevant, as the test has also been found to be suitable for PLWH receiving a tenofovir alafenamide (TAF)-based regimen [32], and oral regimens containing TDF of TAF may be more suitable or preferred over long-acting injectable regimens for many PLWH [33].

This study had some limitations. First, we conducted IDIs with PLWH while their six-month POC viral load test was processing, so our sample did not include any PLWH with viremia at the six-month visit or who were lost to follow-up. However, to ensure our sample included PLWH who had experienced adherence challenges during the study, our sampling schema was defined *a priori* to purposively include PLWH who had received at least one undetectable POC urine tenofovir test result. Second, participants’ responses in this qualitative study may also have been impacted by social desirability bias, as participants may have been more likely to speak favorably about their experience with the intervention. To prevent social desirability biases in this qualitative study, PLWH were interviewed by members of the research team who had not previously had interaction with the study participants, and healthcare providers were interviewed by a member of the research team who didn’t have a supervisory role in the study or within the organization. Third, while the study population is generally reflective of the population of adult PLWH in South Africa and all other procedures implemented in the study were SOC, this trial is being conducted by a clinical research team in research clinics, so our findings may not be generalizable to other settings.

## CONCLUSIONS

Overall, monthly POC urine tenofovir testing in the first five months of treatment was highly acceptable to PLWH who recently initiated ART and to healthcare providers. Perceptions of a urine tenofovir assay to measure adherence were overwhelmingly positive among PLWH and healthcare providers, with very few concerns and no perceived negative effects. In particular, participants in this qualitative study perceived the urine tenofovir assay to have an overall positive impact on adherence, engagement in care, and overall health and wellbeing. When coupled with positive feedback and messaging from providers, a monthly POC urine tenofovir testing intervention may be an effective strategy for motivating optimal adherence for PLWH initiating ART and for facilitating open, honest communications about ART adherence challenges during the course of care.

## Data Availability

All data produced in the present study are available upon reasonable request to the authors.

## ACKNOWLEDGMENTS

We would like to thank the STREAM HIV trial participants, and participants and healthcare providers who participated in this qualitative study. We would also like to acknowledge the research staff at the CAPRISA eThekwini CRS and Vulindlela CRS who contributed their time and effort to this research. The STREAM HIV trial was funded by the National Institute of Allergy and Infectious Diseases (AI147752).

