## Supplemental Table 1 for "Acceptability and perspectives on clinic-based urine tenofovir testing for antiretroviral therapy adherence monitoring: qualitative findings from a randomized controlled trial in South Africa"

**Supplemental Table 1. Additional quotes from STREAM HIV in-depth interviews with study participants and healthcare providers**

| **Theme** | **Quotes** |
| --- | --- |
| **Acceptability Constructs** | |
| **Affective Attitude** | |
| *Positive attitude* | *“It made me happy because it meant I was doing something right. And also, I understood when I was starting medication, the sister explained to me what happens, how things are going to go and the reason why the test is done.” (Intervention participant, male, 40-49 years old)*  *“These tests encouraged me because they are confirming the good work that I’m doing well in taking pills, and that’s makes me happy and see that we’re doing things well.” (Intervention participant, male, 40-49 years old)*  *“I liked it because as I have said it’s encouraging me to take my treatment because if I don’t it will show so I feel it’s a good study and maybe others will be encouraged too to take their pills.” (Intervention participant, female, 40-49 years old)* |
| *Neutral attitude* | *“I didn’t have a problem because I was explained to in the beginning, I didn’t have a problem.” (Intervention participant, male, 40-49 years old)* |
| **Burden** | |
| *Convenience* | *“I think it's non-invasive, and it doesn't cause any prolonged waiting times or inconvenience.” (Healthcare provider, female, urban site)* |
| *Effort* | *“I think it's user-friendly. If I can put it in that way. It's easy to read. It’s easy to actually use it.” (Healthcare provider, female, rural site)*  *“I think the nurses are so streamlined in our study that it all happens very quickly, and the participants know they come, they need to give urine. So I think it's a very efficient process, and I think because the nurses are so efficient, and because it doesn't require too much effort, you know, in terms of getting the urine, waiting time, it doesn't really affect the two arms. There’s not much difference.” (Healthcare provider, female, urban site)*  *“The test is…there is nothing like…there’s nothing much you do with the test, so it doesn’t take much of our time. Because we ask the participant to collect the urine, and then they come back with the urine, and then we start doing the test. Then you wait for like 5 minutes to give results. There’s nothing much like we do in between. So we wait for the results. You do the test, and then we wait for the results, and then you disclose, and then we can continue with whatever else we’re doing. It doesn’t take much or like interfere with something else that we’re doing now.” (Healthcare provider, female, urban site)* |
| **Opportunity Costs** | |
| *Anxiety, discomfort, embarrassment or fear* | *“You get nervous even if you do things the right way and you think the test might say something opposite you know. It is like waiting for the test results at school, so seeing one line really made me feel happy that yah I am doing okay.” (Intervention participant, male, 50-59 years old)*  *“Yeah, at first I think I was very worried, but as time went I saw that it was helping me to continue staying healthy…* *I was worried… You know that when it’s your first-time taking medication sometimes you feel down and it’s like I was not ready the first days, but as time goes on the test it was helping me to continue.” (Intervention participant, female, 30-39 years old)*  *“I was scared because in my first month I used to forget the pills, and I’d go to bed without taking it and it was often so, I didn’t know if it was going to show or not.” (Intervention participant, female, 30-39 years old)*  *“I always felt comfortable because I thought it was a good way that encourages me to take my treatment and it kept me fearful that they’d find out if I’m not taking pills. So it is better to take them so that if they say they’re not found I would ask how if I know I take them all the time.” (Intervention participant, female, 40-49 years old)*  *“I think most would choose the one that doesn't use the test. I think the reasons would be feeling embarrassed carrying a class around with urine. Taking a pee all the time, always going to the toilet. Going to take a pee every time you go to the clinic, they might view that as a huge task.” (Intervention participant, female, 20-29 years old)* |
| **Perceived Effectiveness (and Secondary Benefits)** | |
| *Adherence* | *“…maybe the thoughts of taking my pills on time stays on my mind because I’ll be tested at the clinic. It’s something that’s always on my mind, and I’ll confirm that too when I get to the clinic.” (Intervention participant, female, 40-49 years old)*  *“It’s good. I think it will help others because most people are dodging because there’s nothing to expose them because they are only asked questions like if you take pills correctly and they’ll say yes, but knowing very well that you aren’t taking it, you’ll only be seen by defaulting. But if there’s something to check you, you’d be careful to take it because no one wants to be caught doing bad things, so you’d be able to do the right thing because something will tell you that you didn’t do well here.” (Intervention participant, male, 40-49 years old)*  *“To me personally it doesn’t make much difference because I know I’ve been taking pills correctly, maybe it can make a difference to someone who sometimes cheats on taking pills. To me it doesn’t, I’m willingly taking pills because I want to.” (Intervention participant, male, 40-49 years old)*  *“I think I would prefer the test to be used because it will encourage me to take it, because it’s not easy to take them to be honest. Taking pills every day is a story. So, for me it helped me because I thought it will be hard on me, but these tests motivated me because I know it will show.” (Intervention participant, female, 30-39 years old)*  *“I think that there would be a difference [if there was no adherence testing], because if you’re taking this [test], it makes you take your medication. And if you’re going for your next appointment, you feel that you have to make your nurses proud and to show them that they are teaching me something make my life easy. If it wasn’t for this test I don’t think people would be taking their medication. It’s very hard.” (Intervention participant, female, 30-39 years old)*  *“These tests encouraged me because they are confirming the good work that I’m doing well in taking pills, and that makes me happy and see that we’re doing things well.” (Intervention participant, male, 40-49 years old)*  *“Yes, it can have an impact on people ensuring that they take their pills well because they’d know that when they get to clinic they’d be asked to pee, and it will show that the pill is not in their urine.” (Intervention participant, female, 30-39 years old)*  *“…this test is just encouraging on its own, just by knowing that when you come you’ll have to undergo this test and if I don’t take my treatment the test will prove that. Meaning I’ll be exposed that I’m not doing well, so it is a motivation on its own.” (Intervention participant, male, 40-49 years old)*  *“My only concern is it doesn't sort of give you a long-term view of their adherence because, you know, as long as they've been taking within those few days prior to their visit, and perhaps if they know they've got a visit coming up, they might be more compliant or adherent. Because now, in the back of their mind, they're thinking that, ‘oh, I’ve got a clinic visit next week. Thinking about visits, I should be better with my treatment. Let me start taking my treatment.’ Perhaps some of the longer times where you're away from the clinic that you're less adherent without contact.” (Healthcare provider, female, urban site)*  *“I think it'd actually have a positive outcome [if clinics started using this for everyone] because it would actually encourage them not to miss any dose. So I think it'll improve their adherence.” (Healthcare provider, female, rural site)*  *“There was no problem with it really and I found that it helps to motivate you to take your pills because you know that there is something that will show if you are not taking them. So, makes it easier for you to remember to take the pill because you have just started the treatment and sometimes, we have doubts that should you really take the pill or not but then you get a reminder that you should take them because something will tell on you if you don’t take them.” (Healthcare provider, male, urban site)* |
| *Identifying adherence issues* | *“I liked it, the way I perceive things, is that when something happens it must happen while you are looking at it, so that you can be enthusiastic as the person who is seeking help. But if you know that they don’t check it at that specific time or you don’t know where they are taking it and leave it at that…It is your life, so you need to know how everything is going now that they have tested you. So that they can also be enthusiastic because they can also see that this person cares about their life, therefore let’s keep doing as we are supposed to.” (Intervention participant, female, 30-39 years old)*  *“I think also doctors will benefit. I think because hearing it from someone who self-reports who you can also see that their health is getting worse instead of getting better can be a challenge but if you have the test, you will know if you are dealing with someone who is taking their treatment or someone who has other underlying issues.” (Intervention participant, female, 30-39 years old)*  *“In life it is very nice to know where you stand with your health. It is not nice collecting medication and not getting tested. At the end of the day everything should be tested, even the car is tested. That helps and even encourages you to think you are doing the right thing.” (Intervention participant, male, 50-59 years old)*  *“If I wasn’t using the test, if maybe they just got the information and maybe not ask me the questions, they usually ask about me taking my pills without a record to prove if the pill was actually active in my blood. I’m an honest person so I'm always honest about my treatment, if I was a dishonest person there wouldn’t be a relationship between me and the person assisting me. And also, I’d question how sure they are about my adherence to the treatment because I was just saying it with my mouth without any proof, I could have said anything. And also, I'd think that maybe they didn’t know what they're doing cause how do you prove that I'm really taking them or if they just give it to me and I just throw them away.” (Intervention participant, female, 20-29 years old)*  *“I think it just gives you a better picture of adherence so that you know how to better support and counsel the patient or participant. And I think if you do that early on, so it would just help establish them for a good, lifelong journey with their ART.” (Healthcare provider, female, urban site)*  *“It's something that would help you in terms of monitoring your participants and tracking their progress and whether their treatment is actually effective or not.” (Healthcare provider, female, urban site)*  *“So from my experience with patients, more often than not, they are not entirely truthful when it comes to it adherence. So, as much as it is about the patient, they’re often not the most reliable with this. And we try to, personally even, veer towards more objective measures. So you know, you look at pill counts, you look at viral loads, you look at a urine-based tenofovir test as a more objective marker of adherence compared to what the perception of what the patient is trying to convey. I’ve never…I’ve had very few patients who just came and told me that they’re struggling to take tablets. But when you’ve got the high viral load, or lack of tenofovir in the urine, then the discussion is a bit different. Because then I’m more targeted to say, “You know what, based on the test it looks like you might be struggling to take your doses every day. What’s been your experience? What are the challenges?” And then they do open up. But generally speaking, if you ask them, “How is everything going?” “No, I’m taking my pills every day as I’m supposed to. No issues, no challenges.” So I think it is useful in that way to kind of direct and be a bit more objective.” (Healthcare provider, male, urban site)* |
| *Self-report of adherence* | *“I’d sometimes lie that I took pills well knowing that I did not and find out when the test is done.” (Intervention participant, female, 30-39 years old)*  *“I think it will be for the best if the nurse is told by the test that I have been or not taking the pills because with me I will not really tell the nurse the truth sometimes and she will not be sure herself if I am being honest or not.” (Intervention participant, male, 30-39 years old)*  *“It can have a big role because people will know that if they aren’t taking pills, results will show that they’re not, so it will have to be found out what is the problem and will not be able to deny as the detector is going to encourage people as we know some people would see them fit and well and decide to default but then if tests like this will keep on encouraging as results will tell every time you see a nurse.” (Intervention participant, male, 40-49 years old)*  *“They’re always disappointed [if they get a negative test result]. It always looks like there’s something wrong with the test. But then, they’ll tell you that they haven’t been taking treatment. But then still, they’re like disappointed. You know? They wanted it to be positive. But then they’ll tell you the truth that, ‘No, I haven’t been taking my treatment.’ Maybe ‘I’ve come back at 10:00’ or ‘I’m working at the restaurant now’ or ‘I’ve been traveling with work.’ But still, not look happy with the results.” (Healthcare provider, female, urban site)* |
| *Client-provider relationship and communications* | *“The results have good impact…because we have a conversation, they ask questions, are patient with you. I didn’t feel hesitant.” (Intervention participant, female, 30-39 years old)*  *“It played a big role in maintaining our relationship with the nurses not that I was doing all this for them, but it was encouraging me to carry on so that the nurses will be happy as well, because every time I come here, I get tested, it also encourages me to continue to take my pills well.” (Intervention participant, male, 20-29 years old)*  *“With most of our participants, they are quite adhering. So I would actually praise them for taking their medication and adhering to their medication. So it would be more of a congrats and well done kind of approach.” (Healthcare provider, female, rural site)* |
| **Preferences and Willingness to Use** | |
| *Preferences* | *“I would choose the test because the test really ensures, because verbally you can say you take your treatment well, while knowing very well that you don't. But there is nothing a nurse could do, don't just believe what you're telling them with your mouth.” (Intervention participant, female, 20-29 years old)*  *“I would prefer using the adherence test to make sure that my relationship with the health provider is honest.” (Intervention participant, female, 20-29 years old)*  *“I think it will be for the best if the nurse is told by the test that I have been or not taking the pills because with me I will not really tell the nurse the truth sometimes and she will not be sure herself if I am being honest or not. Sometimes I will say that I have been taking them while I wasn’t. So I prefer the test.” (Intervention participant, male, 30-39 years old)*  *“I’d choose doing tests because even if I can report honestly but not everyone will do that and nurses wouldn’t be able to prove the truth of what is reported.” (Intervention participant, male, 40-49 years old)*  *“That would depend on time I have, but this this thing doesn’t take even five minutes, so if I had to choose… I don’t care about anything because it doesn’t take time to do this thing, even though I take these pills correctly. I don’t have to prove to anyone, but if you want to see for yourself, it’s fine I won’t say anything. But I cannot just volunteer to take it just to see what it will say because I know I take my pills well.” (Intervention participant, male, 40-49 years old)*  *“I think I would do the test. I think because this one would prove. They will see the results. It will prove that I am actually taking the medication because just saying that oh I am talking the medication it won’t prove anything by telling.” (Intervention participant, female, 30-39 years old)*  *“Personally, [I prefer] the test for me. It’s because doing it set me free. Because sometimes I get there and don’t feel like talking. But if I know I will do the test then I’ll know that yeah I can be free and believe it. But as for me I am good with it.” (Intervention participant, female, 20-29 years old)* |
| *Willingness to use POC urine tenofovir test* | *“[I would be] 100% [willing]. Because since we started, because before we started STREAM, I’d never heard of this test. But then, since we’ve started, in seeing the response from participants as well, it looks like a good thing to do, especially for them, the participants. So I think doing this for everyone would also benefit both the clinic or the department and also participants.” (Healthcare provider, female, urban site)* |
| **Appropriateness** | |
| Value and usefulness | *“I saw it simple and right for a person who’s not informed of medical things, though I don’t know more the science behind it showing results based on what nurses and doctors wish to see. But it makes it easy to see if you are doing well or not and to have something practical thing happening in front of you, and you get to be explained to as well even before conducting the test.” (Intervention participant, male, 40-49 years old)*  *“I would love [if all HIV clinics began using adherence testing for all people taking ARVs] because I love to see people well. I have seen in some other clinics you`ll find that a person will be given medication without checking if they use it or not. So, if all clinics can use this test even the number of people that forget to take their medication will decrease and those who get sick from defaulting as well, so it will be a good thing if this test would be available at other clinics as well.” (Intervention participant, male, 20-29 years old)* |
| **Feasibility** | |
| Barriers to implementation | *“It should be used. It should be used. It’s not a difficult test to perform. It’s not a difficult test to interpret either. So I mean, it just means that people have to be willing to do it. Sort of like with urine, TB urine LAM. You know, it’s there, but not everyone is actually using it even though it’s available. And I think this would be the same.” (Healthcare provider, female, urban site)*  *“So I don't know the financial aspect of it, and I’ve worked in both public and private healthcare. So I finished my community service, and then I moved across to private healthcare. And if there’s anything I have learned, most of the decisions made in this country are guided by finance. Not really what’s actually beneficial to patients or what’s the best care for patients. So they try to do what they can do. I mean, it’s an overburdened system. So I don't know what the barriers are like in terms of cost. I don't know what the test kit costs. So I think that would be one of the biggest hurdles. If it were affordable, then I’m sure they would be able to roll it out. But if it's going to be very expensive, then it may not be a feasible thing, considering the amount of HIV we treat in this country and the size of the population in this country.” (Healthcare provider, female, urban site)*  *“I think training as well needs to be done appropriately. I mean a simple thing like screening for TB has become such an issue next door, you know. It's something that should be your bread and butter if you're treating HIV patients. And it’s not being done effectively next door. So if we do roll this out, the training needs to be done appropriately.” (Healthcare provider, female, urban site)* |
